# Response of active trachoma during modified antibiotic mass drug administration in four districts of northern Tanzania, 2022-2023: Results from repeated cross-sectional sentinel site monitoring using serological and molecular testing

**DOI:** 10.1101/2025.06.03.25328347

**Authors:** William E. Oswald, Mabula Kasubi, George Kabona, Molly W. Adams, Alistidia Simon, Ambakisye Mhiche, Veronica Kabona, Donal Bisanzio, Sarah Boyd, Ana Bakhtiari, Cristina Jimenez, Emma M. Harding-Esch, Richard Reithinger, Upendo J. Mwingira, Rebecca M. Flueckiger, Clarer Jones, Jeremiah M. Ngondi

## Abstract

**Background:** Since the first azithromycin mass drug administration (MDA) in 1999, Tanzania has made remarkable progress towards trachoma elimination. Yet several districts have seen prevalence of trachomatous inflammation—follicular (TF) in children aged 1–9 years remain or rebound ≥5%. From 2022, Tanzania modified MDA implementation in these districts to either more frequent than annual (MFTA) or two additional annual MDA. Conducted in four districts receiving MFTA MDA (Longido and Ngorongoro) or annual MDA (Monduli and Simanjiro), our sentinel site monitoring study aimed to: 1) estimate TF prevalence, anti-Pgp3 serology measures, and ocular *Chlamydia trachomatis* (Ct) infection prevalence among children aged 1– 9 years; 2) characterize ocular Ct infection prevalence over time; and, 3) identify factors associated with ocular Ct infection and increasing community Ct infection prevalence.

**Methods:** Ten sentinel sites, with the highest prevalence of active trachoma, were selected per district. At each site, during three monitoring rounds just before MDA in June 2022, January 2023, and October 2023, 50 children aged 1–9 years were examined for trachoma clinical signs and had conjunctival swab and dried blood spot samples taken.

**Results:** Though ocular Ct infection prevalence declined, there was no change in TF prevalence, Pgp3 seroprevalence, or Pgp3 seroconversion during follow-up. Younger age, female gender, and more time required to collect water were associated with higher infection prevalence, and greater distance to a health facility was associated with increasing community infection prevalence.

**Discussion:** Findings support shift from annual to MFTA MDA in Monduli, continued MFTA MDA in Longido and Ngorongoro, and “Wait and Watch” in Simanjiro. Annual monitoring and environmental improvement will be valuable alongside further investigation of relationships between water and sanitation conditions and ocular Ct infection in this setting. Prevalence surveys with additional molecular and serological testing are recommended in the four districts at the end of the MDA cycle.

**What is already known on this topic:**

- Persistent and recrudescent active trachoma is recognized as a late-stage challenge that prevents several countries from attaining elimination of trachoma as a public health problem.
- Evidence is lacking on how to optimally modify and monitor implementation of the surgery, antibiotics, facial cleanliness, and environmental change (SAFE) strategy in settings with persistent and recrudescent active trachoma.

**What this study adds:**

- We selected sentinel sites with greatest likelihood of trachoma transmission in districts with persistent and recrudescent active trachoma to monitor changes over time of clinical, serological, and molecular measures of trachoma during modified mass drug administration (MDA).
- We found fewer ocular *Chlamydia trachomatis* infections following MDA, though TF and serological measures were slow to respond to intervention.
- We observed an association between water availability and individual infection and distance to a health facility with increasing community infection prevalence.

**How this study might affect research, practice, or policy:**

- Our study shows that enhanced monitoring using molecular and serological measures is useful for tailoring implementation of control interventions, like MDA.
- Sentinel site monitoring could be undertaken to help disease control programs decide whether to undertake substantive trachoma prevalence surveys and used for sero-surveillance for trachoma, once elimination has been validated.

## Introduction

Trachoma, caused by ocular variants of *Chlamydia trachomatis* (Ct), remains the world’s leading infectious cause of blindness. The World Health Organization (WHO) estimated in 2024 that, globally, 103.2 million people live in areas endemic for trachoma, and 1.5 million people with trachomatous trichiasis (TT) need surgery to avoid trachomatous blindness in 39 countries.^1^ WHO recommends the use of the SAFE (Surgery, Antibiotics, Facial cleanliness, and Environmental improvement) strategy for trachoma elimination, with the goal of eliminating trachoma as a public health problem by 2030.^2^ Surgery is performed to treat TT, aiming to prevent the progression to blindness in TT-affected individuals. Antibiotic mass drug administration (MDA) with azithromycin is implemented at population-level to clear infections and interrupt transmission; as per 2006 WHO guidelines,^3^ MDA should be implemented when trachomatous inflammation—follicular (TF) prevalence is ≥10% in children aged 1–9 years. Additionally, promoting facial cleanliness and improving access to water and sanitation are crucial for limiting transmission of ocular Ct infection.^3^ WHO recommends population-based prevalence surveys to measure TF among children aged 1–9 years and TT unknown to the health system among adults aged ≥15 years.^4^ A key metric defining trachoma elimination is having a TF prevalence at evaluation unit (EU) level (i.e., a district or division encompassing 100,000–250,000 people) of <5% in children aged 1–9 years.^5^ Following 3–5 years of SAFE implementation, trachoma impact surveys (TIS) are implemented at least six months after the last MDA, and trachoma surveillance surveys (TSS) are implemented two years after a TIS has reported TF prevalence <5% to assess re-emergence.^6^

Tanzania has been implementing the SAFE strategy since 1999, initially on a pilot basis in a few districts. Following baseline surveys between 2004 and 2006 in 50 districts considered most at risk for trachoma^7^, implementation of SAFE was expanded to cover a total of 43 districts that had TF prevalence ≥10%. In 2012 to 2014, a further 31 districts were mapped as part of the Global Trachoma Mapping Project, of which two had TF prevalence ≥5%, warranting MDA.^8,9^ Thus, as of 2022, and accounting for re-districting of administrative areas, there was a total of 69 districts known to be endemic for TF—since then, the Tanzanian Ministry of Health (MOH) has undertaken TIS in 69 and TSS in 65 districts, following SAFE implementation.

Despite implementation of SAFE for over two decades, a total of 10 EUs in nine districts had not attained or maintained the TF elimination threshold by 2021. These EUs were classified as having either persistent active trachoma, where TF prevalence remains ≥5% as measured during two or more TIS, or recrudescent active trachoma, where TF prevalence increases to ≥5% during TSS.^10^ Four of these ten EUs, the districts of Longido, Monduli, Ngorongoro, and Simanjiro, are located in northern Tanzania and primarily populated by the Maasai ethnic group, traditionally nomadic pastoralists who occupy remote settlements. A possible explanation for the burden of disease observed amongst this population is that migrant Maasai are being missed by disease control and elimination activities.^11^ Since 2004, these four districts had each received 8–11 rounds of MDA (https://atlas.trachomadata.org/), but the proportion of the population treated was often inadequate (<80%^12^) in earlier rounds. The MOH conducted TIS in Longido and Simanjiro in 2021 and in Ngorongoro in 2022, while a TSS was done in Monduli in 2021. TF prevalence was >5% in all districts, indicating persistent active trachoma in Longido, Ngorongoro, and Simanjiro and recrudescent active trachoma in Monduli. A recent WHO informal consultation on trachoma elimination endgame challenges in 2021 recommended countries to implement, in addition to TF grading, modified MDA strategies accompanied by testing for ocular Ct infection and Pgp3 serology.^10^ Tanzania subsequently modified its MDA implementation, with annual MDA being extended from one to three years in all nine districts, including Monduli and Simanjiro, and, in Longido and Ngorongoro, adopting more frequent than annual (MFTA) biannual MDA for a period of 3 years.

Based on the WHO informal consultation on enhanced monitoring,^10^ in these four out of nine districts with persistent or recrudescent active trachoma, receiving either annual or MFTA MDA, we conducted sentinel site monitoring with additional serological and molecular testing to: 1) estimate district TF prevalence, seroprevalence of anti-Pgp3 antibodies, anti-Pgp3 antibody seroconversion rate (SCR), and ocular Ct infection prevalence among children aged 1–9 years at three monitoring rounds; 2) describe changes in ocular Ct infection prevalence over time in communities receiving annual or MFTA MDA; and 3) identify factors associated with ocular Ct infection and possible drivers of increasing community Ct infection prevalence.

## Methods

### Study sites and sampling procedure

In each district, we purposively selected ten clusters with the highest proportion of children aged 1–9 years with TF from the most recent prevalence survey as sentinel sites for repeated cross-sectional monitoring (Figure 1). We conducted a power analysis to determine sample size for a proportion comparison, specifying power (1 - β) of 0.80 and α = 0.05 and setting expected prevalence to be 2% (the lowest prevalence expected after MDAs) for a superiority test with a margin threshold set to 4%. Based on this analysis and accounting for 20% non-response, we estimated that at least 300 children per district were required for each survey round. We assumed a survey team could sample 50 children per day, resulting in a minimum of six sentinel sites per district. Finally, to increase the power to detect differences in Ct infection between the annual and MFTA districts at monitoring round 3, we increased the number of sites to 10 per district.

**Figure 1.**
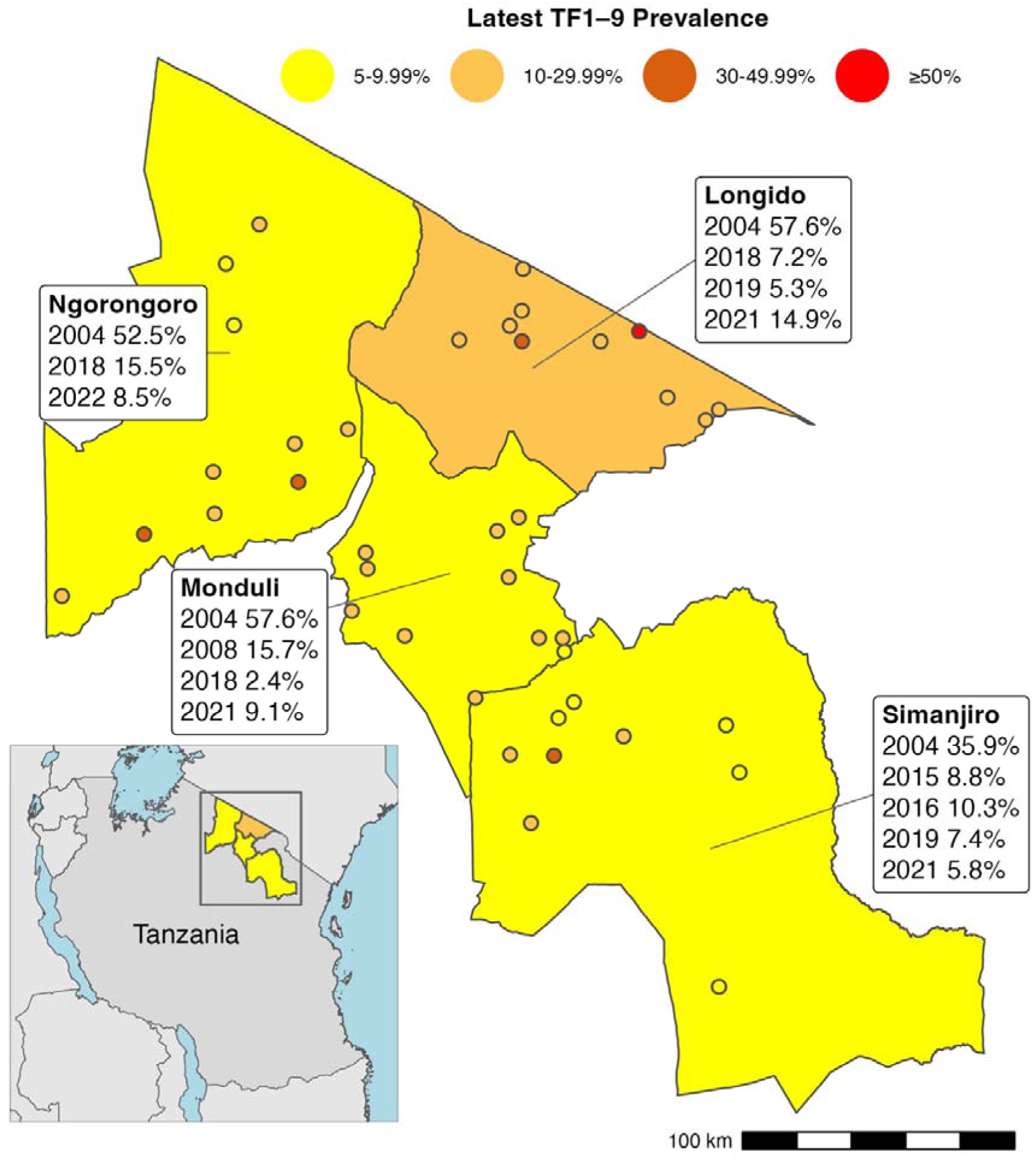
Sentinel site locations and historic prevalence of trachomatous inflammation—follicular (TF) in children aged 1–9 years (TF1–9) of districts included in sentinel site monitoring in Tanzania in 2022-2023.

We conducted cross-sectional monitoring surveys in all 40 sentinel sites in June 2022 (Round 1), January 2023 (Round 2), and October 2023 (Round 3), prior to MDA in all four districts in July 2022, in Longido and Ngorongoro in April 2023, and in all four districts in November 2023. A study timeline is provided in Supplementary Information (Figure S1). At each sentinel site and monitoring round, approximately 20 households were targeted using the compact segment method;^5^ households were surveyed until conjunctival swab and dried blood spot (DBS) samples were collected from 50 children aged 1–9 years.

### Data collection

All survey teams were trained and certified using Tropical Data methods. Four-person data collection teams consisted of a grader (certified to accurately grade TF and TT according to the WHO simplified grading system),^13^ a recorder (certified to accurately record household questionnaire data and clinical examination results using the Tropical Data smartphone application),^13^ a phlebotomist, and a tubing assistant (certified in sample collection and management). We employed eight teams in rounds 1 and 2, completing data collection in five days, and six teams in round 3, completing data collection in seven days.

We collected all data with standardized questionnaires on smartphones with Android operating systems using the Tropical Data (https://www.tropicaldata.org/) application. The Tropical Data questionnaire collected household information on access to water, sanitation, and hygiene facilities, GPS coordinates, and individuals’ gender and age in years. For analysis, reported water sources were classified as improved or not,^14^ and reported time to collect water was classified to be ≤30 or >30 minutes. We assigned a unique number, using a barcoded sticker, to each individual that was scanned within the questionnaire to allow linkage of conjunctival swab and DBS samples with individual data. We used an additional household questionnaire, using the Secure Data Kit (https://www.securedatakit.com) application in round 1 and then Tropical Data for rounds 2 and 3, to collect information on reported household or individual migration in the previous year and the reason for migration.

### Clinical assessment and collection of conjunctival swabs and dried blood spots

Trachoma graders conducted eye examination using a 2.5× magnifying binocular loupe, torch, and follicle size guides.^13,15–17^ We examined all children aged 1–9 years in selected households with parental consent and assent, if applicable, for TF, trachomatous inflammation—intense (TI), trichiasis (upper and lower eyelid, separately), and trachomatous scarring (TS) among those with trichiasis. With the left upper eyelid everted, the grader firmly swabbed the tarsal conjunctiva, using a sterile swab, in a horizontal motion three times, rotating the swab 120° with each motion. The swab shaft was snapped by the grader to fit the swab into a GeneXpert Swab Transport Reagent tube (Cepheid, Sunnyvale, CA) held by the tubing assistant. Samples were immediately placed in a cooler with ice packs for transport. The grader and tubing assistant cleaned their hands with alcohol gel after each examination. The phlebotomist pricked a finger and collected blood onto filter paper (TropBio Pty Ltd., Townsville, Queensland, Australia) with 6 circular extensions calibrated to absorb 10 μL of blood. Filter papers with blood spots were placed in a covered bucket with capacity for 50 samples for transport at ambient temperature. At the district hospital at the end of each day, DBS were packaged individually into sealable plastic bags and stored with desiccant in a larger bag, and swabs and DBS were transferred to a -20°C freezer and stored until the end of data collection.

### Laboratory procedures

De-identified swab and DBS samples were transported to Muhimbili National Hospital in Dar es Salaam, Tanzania and stored at -80°C until processed.

We tested conjunctival swab specimens in pools of 4–5 samples for presence of Ct using the GeneXpert CT/NG Detection Kit (Cepheid, Sunnyvale, CA), a rapid nucleic acid amplification test intended for qualitative detection of Ct DNA. Results of each pool test could be negative, positive, or invalid. If a pool tested positive, the pool was deconstructed and specimens retested individually to identify positive sample(s). If the result of a retest was positive, the sample was considered positive for Ct. Pools with invalid results were retested both as pools and as individual specimens to determine positive or negative results.

We tested DBS samples using a Lateral Flow Assay (LFA) dipstick manufactured at the U.S. Centers for Disease Control and Prevention, using a black latex detector reagent, according to Gwyn et al.:^18^ Pgp3-latex (Expedeon) and Streptavidin-gold (Arista Biologicals) were diluted 1:240 and 1:120, respectively, in PBST (0.3% Tween-20 in phosphate buffer saline) to create a conjugate mastermix. Each DBS was eluted in 60 µL of conjugate mastermix in a well of a flat-bottom 96-well plate, sealed with film, and stored overnight at 4 °C. The next day, Pgp3 LFA dipsticks were added to each well and incubated for 15 min, until all liquid was absorbed. PBST (80 µL) was then added to each well to clear the background caused by hemolyzed red blood cells on the nitrocellulose membrane. Each LFA strip was read as positive, negative, or invalid.

### Sentinel site covariates

For analysis, using data from monitoring round 1, we derived three potential community-level predictors of Ct infection prevalence as the proportion of households that: 1) reported any cattle migration in past year; 2) had any form of sanitation; and 3) had washing water within 30 minutes’ collection time. Using data from routine coverage evaluation surveys conducted after each MDA (Figure S1), we calculated a site measure of “MDA treatment coverage” as the average of the proportion of respondents per site who reported swallowing azithromycin during each MDA, weighted by the number of respondents. We calculated median household latitude and longitude by site across rounds to represent site location. We then used these coordinates to extract community measures of population density in 2020 (1 km)^19^ and distance to Open Street Map major roads in 2023 (100 m)^20^ from publicly available WorldPop datasets. The MOH provided the 2022 listing of health facilities. We restricted this listing to public dispensaries, public health centers, and public hospitals with non-missing GPS coordinates, and used latitude and longitude projected to WGS84/UTM zone 36S (EPSG:32736) to calculate straight-line distance in kilometers to each site. We extracted annual mean temperature in degrees Celsius and annual precipitation in millimeters from WorldClim datasets (1 km),^21^ and we extracted elevation data in meters for each community based on the hole-filled CGIAR Shuttle Radar Topography Mission (90 m) obtained using the *geodata* package.^22^ For analysis, coverage was multiplied by 10, population density was transformed using log_10_, and elevation and precipitation were divided by 100, to facilitate interpretation.

### Data analysis

All analyses were conducted using R version 4.5.0 (https://www.r-project.org).

We estimated TF prevalence, seroprevalence of anti-Pgp3 antibodies, and ocular Ct infection prevalence by district and monitoring round based on mean sentinel site age-adjusted proportions.^23^ Site mean prevalences were first drawn via random selection with replacement, and then the mean of this resulting dataset was bootstrapped with 10,000 iterations. We used the 2.5% and 97.5% centiles of the resulting distribution as 95% confidence intervals (CIs) for each prevalence measure. For each district and monitoring round, we fitted age-dependent seroprevalence curves using cubic splines in generalized additive mixed models with a random intercept for site. We estimated anti-Pgp3 antibody SCR per 100 children per year, among children aged 1–9 and 1–5 years, as the exponentiated intercept from a generalized linear model with binomial error structure and a complementary log–log link, using robust errors to account for survey design.^24,25^

To examine sentinel site Ct infection prevalence over time and predictors of individual infection, we fitted generalized linear mixed effects logistic regression models of individual ocular Ct infection.^26^ Using Akaike Information Criterion (AIC),^27^ we compared a series of nested models: 0) with only random intercept offsets for site; 1) adding, as fixed terms: an indicator for MDA frequency (*i.e.* whether site was in a district assigned to MFTA or annual MDA); follow-up time in months since round 1; and a product term for months of follow-up by MDA frequency to test whether overall trends differed by MDA frequency on the log odds (multiplicative) scale; 2) removing the product term; 3) adding random, site-specific, slope offsets for months of follow-up; 4) a full model, including age in years (centered at the overall mean) and indicators for individual gender, household access to washing water within 30 minutes’ collection time, household access to an improved source of washing water, household access to any form of sanitation; and whether household head reported that the household or a member had migrated to graze cattle within the past year. Following the full model, we fit all possible combinations of these candidate predictors and selected this final model 5 based on the lowest AIC.

From this final model, we used the *marginaleffects* package to: 1) predict Ct infection prevalence by month for each site and overall by MDA frequency (excluding random effects and using mean age and modes for categorical predictors); 2) contrast Ct infection prevalence at the start of follow-up and change in prevalence over the full period of follow-up by MDA frequency; 3) estimate the monthly rate of change in log odds of Ct infection by site; and 4) estimate the average difference in prevalence associated with selected predictors.^28^ We obtained standard errors and confidence intervals for all predictions and contrasts using the Delta method.^28^

We fitted logistic regression models, with robust errors to account for clustering by district, to estimate the association of sentinel site covariates with the odds of having observed a significantly increasing monthly trend for Ct infection.

### Ethics and consent

This study was conducted in accordance with the Declaration of Helsinki. Approval for the activity was provided by the Tanzania National Health Research Ethics Committee (NIMR/HQ/R.8a/Vol. IX/3014). The London School of Hygiene & Tropical Medicine provided ethical approval (16105) for Tropical Data support. Parents or carers for all children provided informed consent, and children aged 6–9 years were asked to provide assent. In addition to household-level consent forms in Swahili signed by parents or carers, we documented consent electronically on the Tropical Data app for every child.

### Patient and public involvement

Study was co-designed and co-led by local and international partners. The study team included representatives from the MOH and local implementing and academic partners. We did not involve patients in the design, conduct, reporting, or dissemination plans for this research. During monitoring round 1, interviews with heads of participating households asked about acceptability of and concerns with swab and DBS collection from children. We disseminated study findings through implementing partners, the MOH, and national and international trachoma technical meetings and scientific conferences.

## Results

We included ten sentinel sites per district. At the first sentinel site monitoring round, we could not access one site in Ngorongoro because of political unrest, and this site was replaced with the eleventh highest TF prevalence cluster from the previous trachoma survey. Out of the total enumerated in each round, round 1 included 1949/2024 (96.3%) children, round 2 included 1991/2019 (98.6%) children, and round 3 included 1961/2007 (97.7%) children aged 1–9 years with complete data (Figure S2).

Participants were balanced across districts by round (Table S1). Of 5901 children included, 3175 (53.8%) were female. Included children were most frequently aged 3 and 4 years in each round. Participants’ household washing water access within 30 minutes’ collection time increased between rounds from 20.2% to 34.8%; households of 3048/5901 (51.7%) included children had an improved source for washing water. Between rounds, participants’ household open defecation ranged from 74.2% (1478/1991) to 91.2% (1788/1961). Of 5901 participants, heads of 1214 participants’ households (20.6%; range between rounds: 9.1–26.8%) reported some migration with cattle in the past year.

Table 1 shows district estimates of prevalence of TF, anti-Pgp3 antibodies, and ocular Ct infection in children aged 1–9 years by monitoring round. Initial TF prevalence was 21.4% (95% CI: 15.5–30.7) in Longido, 17.4% (95% CI: 12.7–24.9) in Monduli, 14.3% (95% CI: 8.7–23.4) in Ngorongoro, and 2.9% (95% CI: 0.7–6.6) in Simanjiro. TF prevalence showed little change over time, and at round 3, estimated TF prevalence in all four districts was ≥5%. We observed no change in district Pgp3 seroprevalence between rounds (Table 1) and little change in age-dependent seroprevalence between rounds (Figure 2). District SCR were also similar between rounds, and all were >3.8 per 100 person-years (Figure 2). At round 3, SCR per 100 person-years among children aged 1–9 years was 13.8 (95% CI: 10.1–18.7) in Longido, 9.4 (95% CI: 6.1–14.5) in Monduli, 17.3 (95% CI: 10.3–29.0) in Ngorongoro, and 5.5 (95% CI: 2.8–10.5) in Simanjiro. SCR among children aged 1–5 years were similar to estimates including older children (Table S2). District ocular Ct infection prevalence estimates decreased in Longido between round 1 to round 2. We observed no other difference in district ocular Ct infection prevalence between rounds. At monitoring round 3 in Simanjiro, we detected 1 case per 1000 people (prevalence=0.1%, 95% CI: 0.0–0.4).

**Figure 2.**
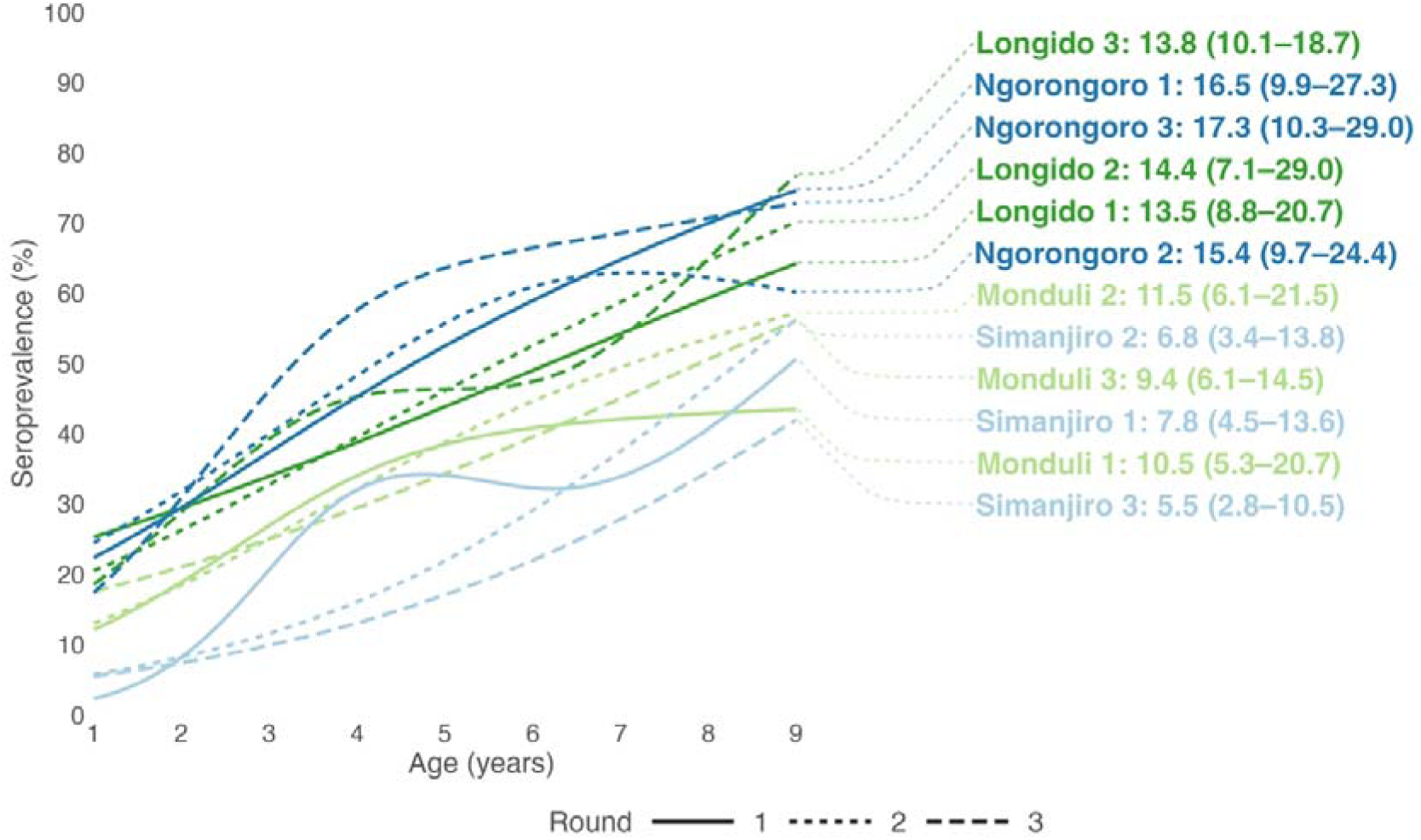
Proportion Pgp3 antibody positive by age in each district and monitoring round. Lines are cubic splines fit to seroprevalence by age. Displayed values are estimated seroconversion rates per 100 person-years among children aged 1**–**9 years with 95% confidence intervals based on robust standard errors.

**Table 1.** Prevalence of trachomatous inflammation—follicular (TF), anti-Pgp3 antibodies, and ocular *Chlamydia trachomatis* (Ct) infection in children aged 1–9 years by monitoring round and district. 95% Confidence Intervals (CI) based on bootstrapped sampling distribution.

| Indicator | Round | Longido<br>Prevalence<br>(95% CI) | Monduli<br>Prevalence<br>(95% CI) | Ngorongoro<br>Prevalence<br>(95% CI) | Simanjiro<br>Prevalence<br>(95% CI) |
| --- | --- | --- | --- | --- | --- |
| TF prevalence | 1 | 21.4 (15.5–30.7) | 17.4 (12.7–24.9) | 14.3 (8.7–23.4) | 2.9 (0.7–6.6) |
|  | 2 | 9.4 (5.2–14.1) | 29.6 (17.7–47.1) | 17.0 (10.2–28.2) | 0.2 (0.0–0.5) |
|  | 3 | 19.4 (10.6–27.0) | 12.7 (7.5–19.8) | 15.3 (11.8–18.9) | 6.5 (3.9–9.0) |
| Pgp3 seroprevalence | 1 | 43.1 (30.4–55.8) | 36.7 (25.6–55.3) | 46.6 (34.5–60.9) | 28.8 (18.8–40.4) |
|  | 2 | 42.2 (27.4–57.3) | 39.7 (28.8–57.8) | 49.0 (40.5–62.5) | 27.2 (17.8–41.0) |
|  | 3 | 44.2 (36.2–51.0) | 34.9 (27.5–46.8) | 51.0 (41.5–66.2) | 23.8 (14.9–35.7) |
| Ct infection prevalence | 1 | 20.7 (9.7–36.5) | 8.9 (4.9–13.7) | 13.6 (3.7–29.1) | 1.2 (0.0–2.9) |
|  | 2 | 5.1 (1.9–9.1) | 4.5 (0.8–10.8) | 7.6 (2.3–17.1) | 0.4 (0.0–1.2) |
|  | 3 | 5.1 (0.6–9.5) | 3.8 (1.1–7.1) | 6.7 (1.4–13.2) | 0.1 (0.0–0.4) |

In 480 households in round 1 with complete data, during interviews, 460 (95.8%) heads of households reported being very comfortable with blood collection and 461 (96.0%) with conjunctival swab collection from their children, and 469/480 (97.7%) would consent to their children’s participation in future testing (Table S3).

Figure 3 shows modelled site-specific and overall mean Ct infection prevalence trends over 16 months of follow-up. Baseline site-specific prevalence varied considerably (SD=1.73, profile 95% CI: 1.28–2.45, model 0, Table S4). A product term allowing rate of change in log odds of Ct infection for each month of follow-up to vary between MDA frequency assignment was not significant (p=0.64, model 1, Table S4). Model 3, excluding this term and including random, site-specific, intercept and slope offsets, had better fit (AIC=2168.62, Table S4). The final selected model, comparing all possible combinations of candidate predictors (model 5), included terms for MDA frequency, months of follow-up, individual age and gender, access to water for hygiene within 30 minutes’ collection time, and any reported cattle migration in the past year (Table S4).

**Figure 3.**
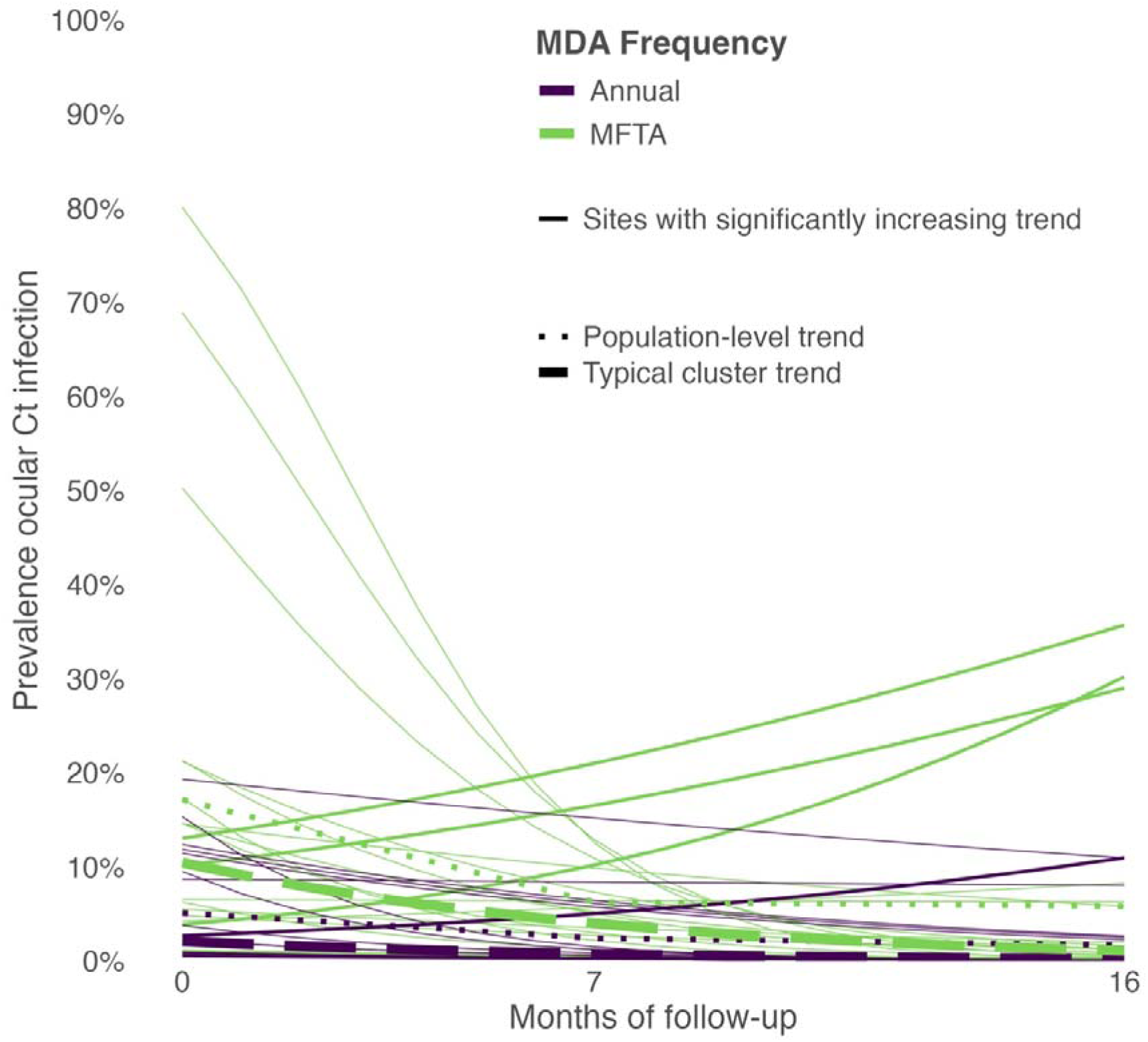
Site-specific and overall trends of predicted ocular *Chlamydia trachomatis* (Ct) infection prevalence over months of follow-up by district assignment to more frequent than annual (MFTA) biannual or annual mass drug administration (MDA), adjusted for age, gender, household water availability for hygiene purposes, and reported migration. Heavy lines indicate sentinel sites with significantly increasing trend.

From this model, predicted prevalence of Ct infection at the start of monitoring was 9.3 percentage points (95% CI: 2.1–16.6) higher among sites in districts assigned to MFTA MDA. During 16 months’ follow-up, predicted prevalence in sites in districts assigned to MFTA MDA declined by 8.7 percentage points (95% CI: 2.4–15.1), while predicted prevalence in sites in districts assigned to annual MDA declined by 1.6 percentage points (95% CI: 0.1–3.1). Each year of age over mean age was associated with a decrease in prevalence of -0.2 (95% CI: -0.3–0.0, p=0.047, *p shown as CI includes 0*). Being female was associated with an increase in prevalence of 1.1 (95% CI: 0.3–1.9). Individuals in households with access to water for hygiene within 30 minutes’ collection time had 1.7 lower prevalence (95% CI: 0.4– 1.3). Household migration was not associated with Ct infection prevalence (Difference = -0.7, 95% CI: -1.7–0.2).

We estimated the change in log odds of Ct infection per month of follow-up for each site and identified sites where this estimate was >0 and its confidence interval did not contain zero. During a period with MDA implementation, we found 4/40 (10.0%) sites with a monthly change in the log odds of Ct infection greater than zero, indicating increasing ocular Ct infection prevalence (Figure 3), 3/4 sites located in districts receiving MFTA MDA (Figure S3). Across all 40 sites, higher baseline ocular Ct infection prevalence decreased odds of observing increasing site Ct infection prevalence (OR 0.20, 95% CI: 0.10–0.38). Adjusting for district, each additional kilometer in distance of the site from a public health facility increased the odds of observing increasing site Ct infection prevalence by 8% (OR 1.08, 95%CI: 1.05– 1.10, Table 2). Higher sanitation coverage and population density were protective against increasing Ct infection, but estimated confidence intervals included one. No other evaluated community predictors showed evidence of association with increasing Ct infection prevalence during follow-up.

**Table 2.**
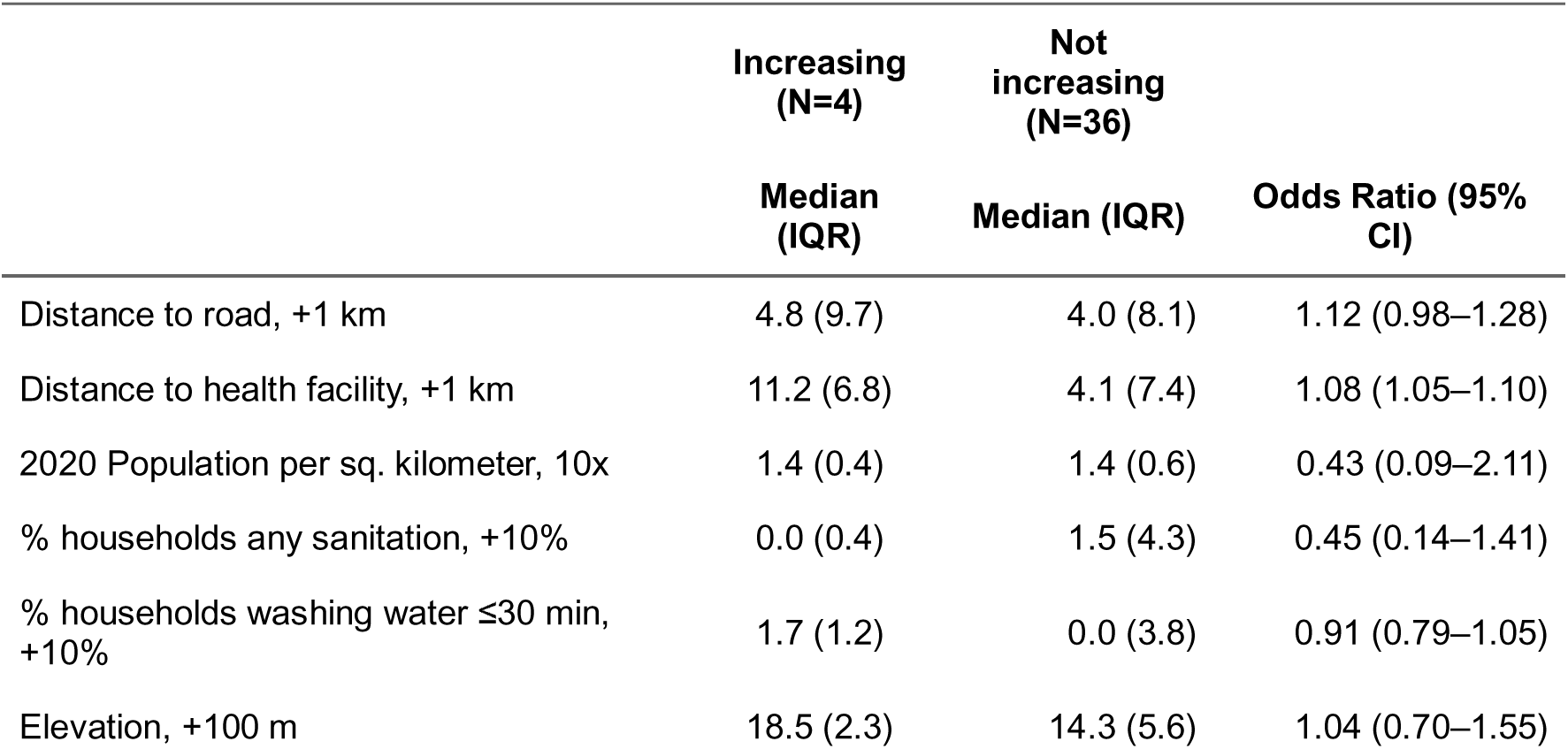
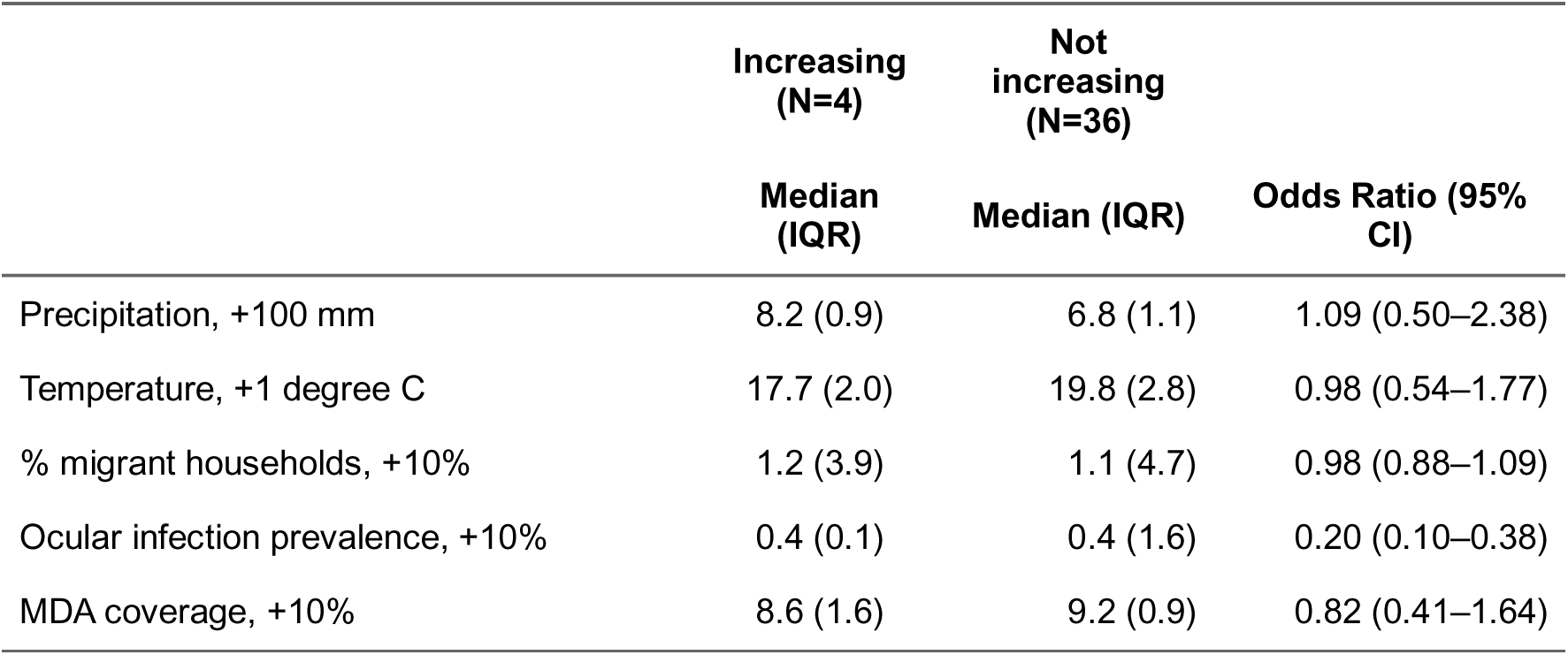
Results from logistic regression models of site (n=40) ocular *Chlamydia trachomatis* (Ct) infection prevalence trend (increasing *versus* not) and candidate predictors. All models adjusted for district with 95% confidence intervals based on robust standard errors.

|  | Increasing<br>(N=4) | Not<br>increasing<br>(N=36) |  |
| --- | --- | --- | --- |
|  | Median<br>(IQR) | Median (IQR) | Odds Ratio (95%<br>CI) |
| Distance to road, +1 km | 4.8 (9.7) | 4.0 (8.1) | 1.12 (0.98–1.28) |
| Distance to health facility, +1 km | 11.2 (6.8) | 4.1 (7.4) | 1.08 (1.05–1.10) |
| 2020 Population per sq. kilometer, 10x | 1.4 (0.4) | 1.4 (0.6) | 0.43 (0.09–2.11) |
| % households any sanitation, +10% | 0.0 (0.4) | 1.5 (4.3) | 0.45 (0.14–1.41) |
| % households washing water ≤30 min,<br>+10% | 1.7 (1.2) | 0.0 (3.8) | 0.91 (0.79–1.05) |
| Elevation, +100 m | 18.5 (2.3) | 14.3 (5.6) | 1.04 (0.70–1.55) |
|  | Median<br>(IQR) | Median (IQR) | Odds Ratio (95%<br>CI) |
| Precipitation, +100 mm | 8.2 (0.9) | 6.8 (1.1) | 1.09 (0.50–2.38) |
| Temperature, +1 degree C | 17.7 (2.0) | 19.8 (2.8) | 0.98 (0.54–1.77) |
| % migrant households, +10% | 1.2 (3.9) | 1.1 (4.7) | 0.98 (0.88–1.09) |
| Ocular infection prevalence, +10% | 0.4 (0.1) | 0.4 (1.6) | 0.20 (0.10–0.38) |
| MDA coverage, +10% | 8.6 (1.6) | 9.2 (0.9) | 0.82 (0.41–1.64) |

## Discussion

We monitored 40 sentinel sites with the highest prevalence of active trachoma in four districts of northern Tanzania during three repeated cross-sectional surveys between June 2022 and October 2023 to characterize changes in TF prevalence, Pgp3 serological measures, and ocular Ct infection prevalence during implementation of MFTA biannual and annual MDA with azithromycin. Findings suggest that community transmission of ocular Ct in Longido, Ngorongoro, and Monduli is ongoing. We identified individual and household predictors of ocular Ct infection. We found four sites where prevalence of Ct infection increased during follow-up despite MDA and found that these sites were located at a greater distance from recorded public health facilities, compared to sites without increasing prevalence.

Molecular detection of ocular Ct infection and measurement of anti-Ct antigen Pgp3 antibodies are recommended for use in settings with persistent and recrudescent transmission,^10^ and trachoma control programs have begun incorporating additional testing in routine surveys.^29–33^ In the current work, we detected no change in district seroprevalence or SCR during 16 months’ follow-up. This observation is expected as only children under one year-old, who were not assessed in the current study, would have been novelly exposed to the lower Force Of Infection (FOI) we might expect to result from increased MDA frequency.^34^ Based on round 3 data; however, SCR estimates, as a measure of FOI, indicate continued community transmission of ocular Ct infection in Longido, Ngorongoro, and Monduli, warranting implementation of biannual MDA. Simanjiro’s SCR of 5.7 (2.7–11.9) among children aged 1–5 years was above recommended thresholds for continued monitoring after halting MDA,^24^ in which case a “wait and watch” approach may be warranted.^35^ The clinical indicator, TF, was also unable to detect change within the time period of the study, and, at round 3, district prevalence estimates were all higher than the 5% threshold recommended for the discontinuation of MDA.^36^ Measurement of ocular Ct infection detected changes over time at the district level and more specifically at the site level. This analysis was strengthened by our testing of individual samples constituting positive pooled samples, which allowed us to examine the association of individual factors and household conditions with ocular Ct infection.

In contrast to studies in Ethiopia, Guinea-Bissau, Niger, and Tanzania that examined individual factors associated with ocular Ct infection,^37–47^ we observed that female children had a higher prevalence than male children, adjusting for age, follow-up month, MDA frequency, and other factors. Similar to our findings, an earlier study in a village in Kongwa, Tanzania, found that among those under-15 years, female children had higher ocular Ct infection prevalence; though in two other sites, prevalence was similar between genders.^48^ Among children, there may be little difference in prevalence of TF between boys and girls, but adult women are known to bear a greater burden of TT than men, likely because of their larger contribution to childcare in most settings.^49^ Previous studies have found a consistent relationship between both household sanitation access and community sanitation coverage with signs of TF and ocular Ct infection.^50–52^ We observed a protective association between household access to any sanitation and individual ocular Ct infection, and higher community sanitation coverage was associated with lower odds of increasing community prevalence. These findings were not statistically significant, however, which may be a result of the overall low sanitation coverage observed. An alternative explanation may be that household sanitation access has a less-pronounced effect in this setting because of residents’ proximity to livestock.

The population in this setting is predominantly Maasai who may practice nomadic pastoralism and occupy characteristic homesteads, called *boma*, that typically consist of one or more cattle corrals surrounded by a ring of familial dwellings.^53^ The bazaar fly, *Musca sorbens*, a mechanical vector for Ct transmission, can breed in animal dung and in numbers that may have limited the observable protection of household sanitation, even where facilities were present and in use. A systematic review of the health risks associated with exposure to animal feces concluded that there was a possible increased risk of trachoma based on examples from Nigeria and Ethiopia, where active trachoma was associated with either living proximity to cattle or presence of animal feces near study households.^54^ In a setting where vector-borne transmission may be common, the availability of water for face-washing may be a more proximal determinant of ocular Ct infection. While a review of water, sanitation, and hygiene conditions and trachoma found no association between distance to water and Ct infection, it included only four studies.^55^ We observed a protective association between water availability for hygiene, indicated by reported presence of a source within 30 minutes’ collection time or less, and individual infection. Though selected to improve model fit, any cattle migration of the whole household or individuals was not associated with individual Ct infection. Similarly, at the site level, we did not observe an association between the proportion of households with reported migration and an increasing prevalence of ocular Ct infection. One hypothesized explanation for the persistent or recrudescent active trachoma in these settings is that migrant Maasai pastoralists are not being reached with treatment during MDA activities. Our findings support the conclusion that recent efforts to align MDA implementation with known migration timings and coordinate cross-border MDA with the Kenya Ministry of Health are successfully reaching and treating these groups.^11^

Treatment coverage following MDA is routinely measured using coverage evaluation surveys.^12^ Our sentinel sites were purposively included in these activities, providing a site-specific measure of coverage after each annual or biannual MDA activity. We did not find an association between the odds of the community having an increasing ocular Ct infection prevalence during follow-up and MDA treatment coverage during the same period, but overall coverage was high in communities with increasing and non-increasing ocular Ct infection (Table 2). Distance of the site from the nearest public health facility was associated with higher odds of increasing Ct infection during follow-up, which may indicate that more remote sites were at greater risk of continued transmission. Other studies have shown that distance to health facility or similar measures of remoteness are predictors of TF.^56^ Additional research is warranted to understand drivers of infection prevalence in remote sites, despite high treatment coverage and increased frequency of MDA.

Our sentinel site monitoring study was strengthened by use of serological and molecular testing. Ct infection prevalence estimates may have underestimated the true burden of ocular Ct infection because only a single eye per individual was tested. Though unlikely to be a large source of misclassification in the included age range, our assays could not distinguish between ocular and urogenital Ct infections. Our conclusions are also limited by use of reported measures at the individual and household level. The generalizability of our findings may be another limitation, as sentinel sites were purposefully selected as ones with highest TF and may not represent typical conditions across each district. However, these sites were considered, and as was shown, to be ones at risk of persistent or recrudescent active trachoma.

In conclusion, our results support a shift to a MFTA MDA strategy in Monduli, continued MFTA treatment in Longido and Ngorongoro, and “Wait and Watch” in Simanjiro. Continued monitoring once per year to inform interventions to reduce community transmission of ocular Ct and strengthening of Facial cleanliness and Environmental improvement (F&E) components of SAFE will be valuable to the Tanzania Neglected Tropical Disease control program. Further research to understand the relationships between water and sanitation conditions with ongoing transmission is needed in these challenging settings and will be facilitated as use of additional testing, particularly molecular detection of ocular Ct infection, becomes more routine. The sentinel site approach may have programmatic applications for monitoring “Wait and Watch” settings, exploring if districts with persistent or recrudescent active trachoma are ready for TSS, and for post-validation sero-surveillance.

## Data Availability

All data produced in the present study are available upon reasonable request to the authors.

## Acknowledgments

We recognize the efforts and contributions of the district health officers, supervisors, and study participants without whom this study would not be possible. We acknowledge the Tanzania Ministry of Health for their support and Dr. Diana Martin and Dr. Sarah Gwyn at the U.S. Centers for Disease Control and Prevention (CDC), whose expertise and advice were invaluable to the success of this activity. Laboratory resources for serological testing at Muhimbili National Hospital were provided by the CDC through an interagency agreement with United States Agency for International Development (USAID).

## Data availability

De-identified participant data are available upon request to the principal investigators.

## Author contributions

MK, GK, RR, UJM, RMF, CJ, DB, JMN contributed to study conception and design. SB, AB, CJ, EMHE reviewed methods and supported training and data collection and management. MK, GK, MWA, AS, AM, VK contributed to the execution of the field work. MK conducted laboratory work. WEO analyzed data and wrote manuscript. All authors reviewed and approved the manuscript.

