## Supplementary tables and figures for "Response of active trachoma during modified antibiotic mass drug administration in four districts of northern Tanzania, 2022-2023: Results from repeated cross-sectional sentinel site monitoring using serological and molecular testing"

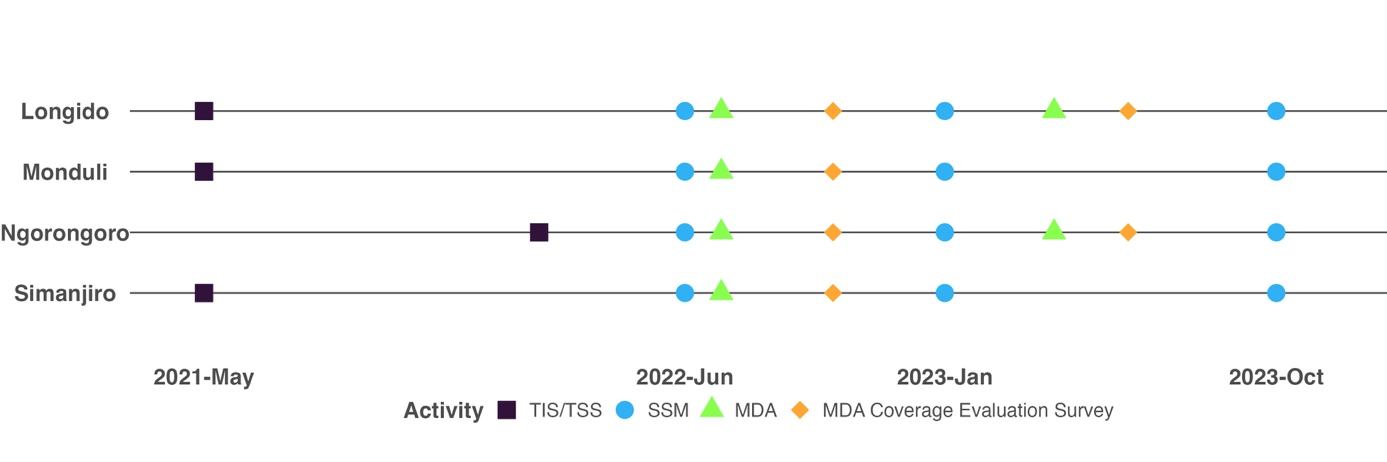

**Figure S1. Dates of data collection activities and mass drug administration**

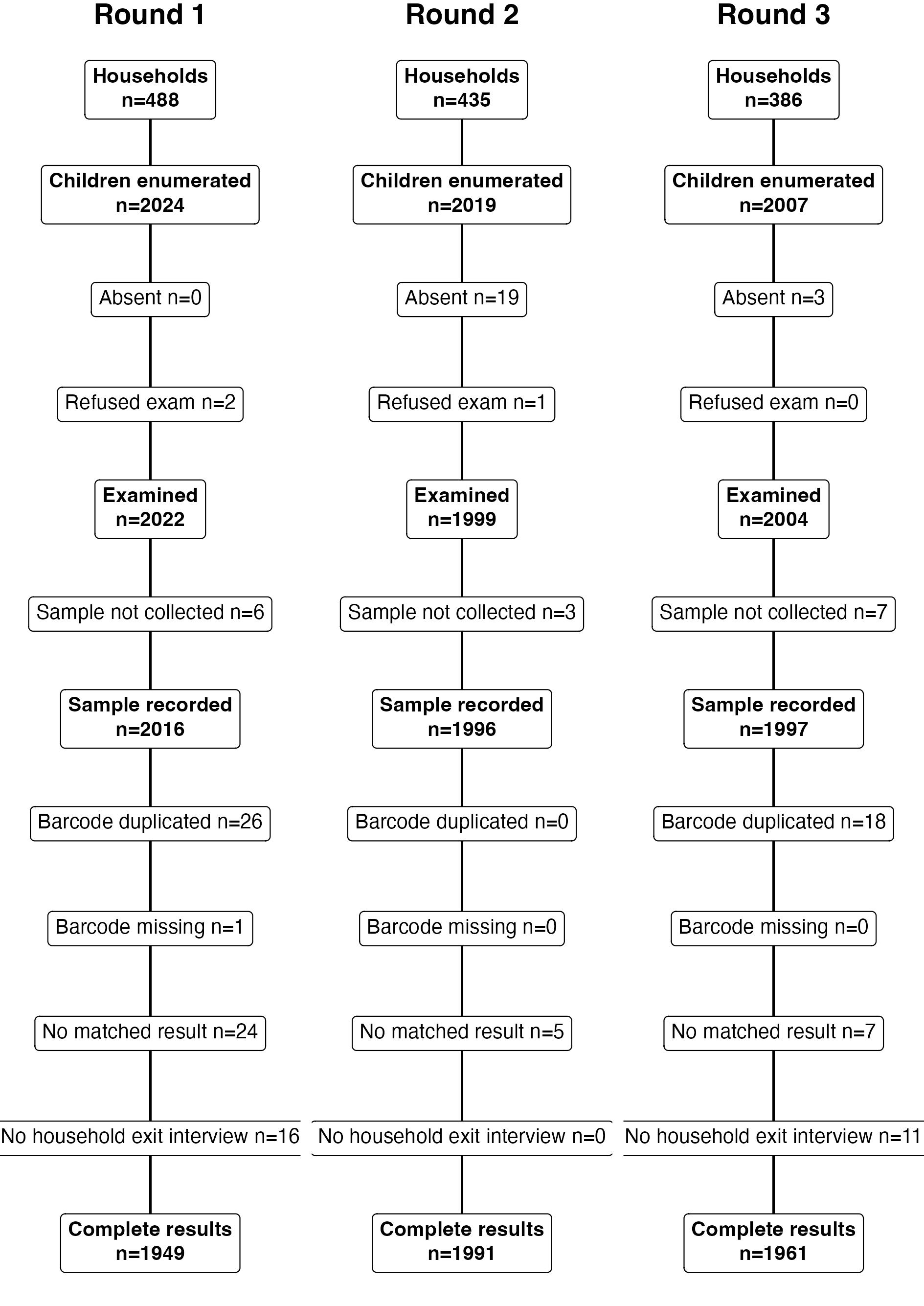

**Figure S2. Numbers of households and individuals included in analysis by round**

**Table S1. Characteristics of participants in three rounds of cross-sectional monitoring in northern Tanzania in 2022 and 2023.**

|  |  | **Round 1 (N=1949)** | **Round 2 (N=1991)** | **Round 3 (N=1961)** | **All (N=5901)** |
| --- | --- | --- | --- | --- | --- |
|  |  | **n (%)** | **n (%)** | **n (%)** | **n (%)** |
| **District** | Longido | 498 (25.6%) | 499 (25.1%) | 488 (24.9%) | 1485 (25.2%) |
|  | Monduli | 480 (24.6%) | 499 (25.1%) | 487 (24.8%) | 1466 (24.8%) |
|  | Ngorongoro | 490 (25.1%) | 496 (24.9%) | 497 (25.3%) | 1483 (25.1%) |
|  | Simanjiro | 481 (24.7%) | 497 (25.0%) | 489 (24.9%) | 1467 (24.9%) |
| **Gender** | Male | 900 (46.2%) | 915 (46.0%) | 911 (46.5%) | 2726 (46.2%) |
|  | Female | 1049 (53.8%) | 1076 (54.0%) | 1050 (53.5%) | 3175 (53.8%) |
| **Age, years** | 1 | 240 (12.3%) | 228 (11.5%) | 168 (8.6%) | 636 (10.8%) |
|  | 2 | 273 (14.0%) | 304 (15.3%) | 265 (13.5%) | 842 (14.3%) |
|  | 3 | 318 (16.3%) | 335 (16.8%) | 281 (14.3%) | 934 (15.8%) |
|  | 4 | 300 (15.4%) | 290 (14.6%) | 328 (16.7%) | 918 (15.6%) |
|  | 5 | 275 (14.1%) | 248 (12.5%) | 248 (12.6%) | 771 (13.1%) |
|  | 6 | 197 (10.1%) | 203 (10.2%) | 184 (9.4%) | 584 (9.9%) |
|  | 7 | 144 (7.4%) | 164 (8.2%) | 167 (8.5%) | 475 (8.0%) |
|  | 8 | 118 (6.1%) | 120 (6.0%) | 163 (8.3%) | 401 (6.8%) |
|  | 9 | 84 (4.3%) | 99 (5.0%) | 157 (8.0%) | 340 (5.8%) |
| **Household improved washing water source** | No | 886 (45.5%) | 967 (48.6%) | 1000 (51.0%) | 2853 (48.3%) |
|  | Yes | 1063 (54.5%) | 1024 (51.4%) | 961 (49.0%) | 3048 (51.7%) |
| **Household washing water source ≤30 min** | No | 1555 (79.8%) | 1437 (72.2%) | 1279 (65.2%) | 4271 (72.4%) |
|  | Yes | 394 (20.2%) | 554 (27.8%) | 682 (34.8%) | 1630 (27.6%) |
| **Household any sanitation access** | No | 1515 (77.7%) | 1478 (74.2%) | 1788 (91.2%) | 4781 (81.0%) |
|  | Yes | 434 (22.3%) | 513 (25.8%) | 173 (8.8%) | 1120 (19.0%) |
| **Household cattle migration in past year** | No | 1447 (74.2%) | 1458 (73.2%) | 1782 (90.9%) | 4687 (79.4%) |
|  | Yes | 502 (25.8%) | 533 (26.8%) | 179 (9.1%) | 1214 (20.6%) |

**Table S2. Pgp3 seroconversion rate (SCR) by district and round for children aged 1–9 and 1–5 years. 95% Confidence Intervals (CI) based on robust standard errors.**

| **District** | **Round** | **1–9 years SCR (95% CI)** | **1–5 years SCR (95% CI)** |
| --- | --- | --- | --- |
| **Longido** | 1 | 13.5 (8.8–20.7) | 14.2 (7.7–26.1) |
|  | 2 | 14.4 (7.1–29.0) | 16.7 (8.8–31.6) |
|  | 3 | 13.8 (10.1–18.7) | 15.3 (10.8–21.7) |
| **Monduli** | 1 | 10.5 (5.3–20.7) | 12.5 (6.3–24.5) |
|  | 2 | 11.5 (6.1–21.5) | 11.8 (6.5–21.4) |
|  | 3 | 9.4 (6.1–14.5) | 10.0 (6.6–15.1) |
| **Ngorongoro** | 1 | 16.5 (9.9–27.3) | 18.2 (11.0–30.0) |
|  | 2 | 15.4 (9.7–24.4) | 18.5 (10.9–31.5) |
|  | 3 | 17.3 (10.3–29.0) | 20.8 (12.0–35.8) |
| **Simanjiro** | 1 | 7.8 (4.5–13.6) | 8.4 (4.8–14.8) |
|  | 2 | 6.8 (3.4–13.8) | 7.2 (3.3–15.9) |
|  | 3 | 5.5 (2.8–10.5) | 5.7 (2.7–11.9) |

**Table S3. Monitoring round 1 participation acceptability results.**

|  |  | **n (%)** |
| --- | --- | --- |
| **How comfortable/satisfied are you with the eye exam your child/children received today?** | Very comfortable | 455 (94.8%) |
|  | A little comfortable | 22 (4.6%) |
|  | A little uncomfortable | 1 (0.2%) |
|  | Very uncomfortable | 2 (0.4%) |
| **How comfortable/satisfied are you with the dried blood spot sample collected from your child/children today?** | Very comfortable | 460 (95.8%) |
|  | A little comfortable | 18 (3.8%) |
|  | A little uncomfortable | 2 (0.4%) |
|  | Very uncomfortable | 0 (0.0%) |
| **How comfortable/satisfied are you with the eye swab collected from your child/children today?** | Very comfortable | 461 (96.0%) |
|  | A little comfortable | 15 (3.1%) |
|  | A little uncomfortable | 3 (0.6%) |
|  | Very uncomfortable | 1 (0.2%) |
| **Was your child/children’s participation in today’s eye examination and sample collection burdensome for your household?** | No | 471 (98.1%) |
|  | Yes | 9 (1.9%) |
| **Would you consent to your children’s participation in a future trachoma health examination and testing like the one they participated in today?** | No | 11 (2.3%) |
|  | Yes | 469 (97.7%) |

**Table S4. Results from generalized linear mixed effect model selection.**

|  |  | **Model 0** | | **Model 1** | | **Model 2** | | **Model 3** | | **Model 4** | | **Model 5** | |
| --- | --- | --- | --- | --- | --- | --- | --- | --- | --- | --- | --- | --- | --- |
|  | **Variable** | **β (95% CI)** | **p** | **β (95% CI)** | **p** | **β (95% CI)** | **p** | **β (95% CI)** | **p** | **β (95% CI)** | **p** | **β (95% CI)** | **p** |
| Fixed effects | Annual MDA |  |  | 0.00 |  | 0.00 |  | 0.00 |  | 0.00 |  | 0.00 |  |
|  | MFTA MDA |  |  | 1.81 (0.75–2.87) | 0.0008 | 1.77 (0.72–2.81) | 0.0009 | 1.79 (0.68–2.90) | 0.0016 | 1.94 (0.85–3.03) | 0.0005 | 1.98 (0.90–3.06) | 0.0003 |
|  | Follow-up (months) |  |  | -0.08 (-0.12–-0.05) | <0.0001 | -0.09 (-0.11–-0.07) | <0.0001 | -0.15 (-0.23–-0.07) | 0.0001 | -0.15 (-0.23–-0.07) | 0.0002 | -0.15 (-0.23–-0.07) | 0.0002 |
|  | MDA * Follow-up |  |  | -0.01 (-0.05–0.03) | 0.64 |  |  |  |  |  |  |  |  |
|  | Mean-centered age (years) |  |  |  |  |  |  |  |  | -0.06 (-0.11–-0.01) | 0.024 | -0.06 (-0.11–-0.01) | 0.024 |
|  | Male gender |  |  |  |  |  |  |  |  | 0.00 |  | 0.00 |  |
|  | Female gender |  |  |  |  |  |  |  |  | 0.42 (0.18–0.66) | 0.0007 | 0.42 (0.18–0.66) | 0.0007 |
|  | Water >30 min |  |  |  |  |  |  |  |  | 0.00 |  | 0.00 |  |
|  | Water ≤30 min |  |  |  |  |  |  |  |  | -0.70 (-1.15–-0.24) | 0.0027 | -0.70 (-1.15–-0.24) | 0.0026 |
|  | Unimproved water source |  |  |  |  |  |  |  |  | 0.00 |  |  |  |
|  | Improved water source |  |  |  |  |  |  |  |  | 0.04 (-0.49–0.57) | 0.89 |  |  |
|  | No household sanitation |  |  |  |  |  |  |  |  | 0.00 |  |  |  |
|  | Any household sanitation |  |  |  |  |  |  |  |  | -0.25 (-0.67–0.17) | 0.25 |  |  |
|  | No migration |  |  |  |  |  |  |  |  | 0.00 |  | 0.00 |  |
|  | Migration |  |  |  |  |  |  |  |  | -0.28 (-0.64–0.09) | 0.14 | -0.28 (-0.64–0.08) | 0.13 |
| Random effects (SD) | Site-specific intercept | 1.73 (1.28–2.45) |  | 1.51 (1.11–2.16) |  | 1.51 (1.12–2.16) |  | 1.70 (1.25–2.40) |  | 1.65 (1.21–2.34) |  | 1.64 (1.20–2.32) |  |
|  | Site-specific follow-up |  |  |  |  |  |  | 0.17 (0.13–0.24) |  | 0.17 (0.13–0.24) |  | 0.17 (0.13–0.24) |  |
| Model fit | AIC | 2470.70 |  | 2371.17 |  | 2369.37 |  | 2168.62 |  | 2152.02 |  | 2149.41 |  |
|  | AUC | 0.50 |  | 0.83 |  | 0.83 |  | 0.88 |  | 0.89 |  | 0.89 |  |

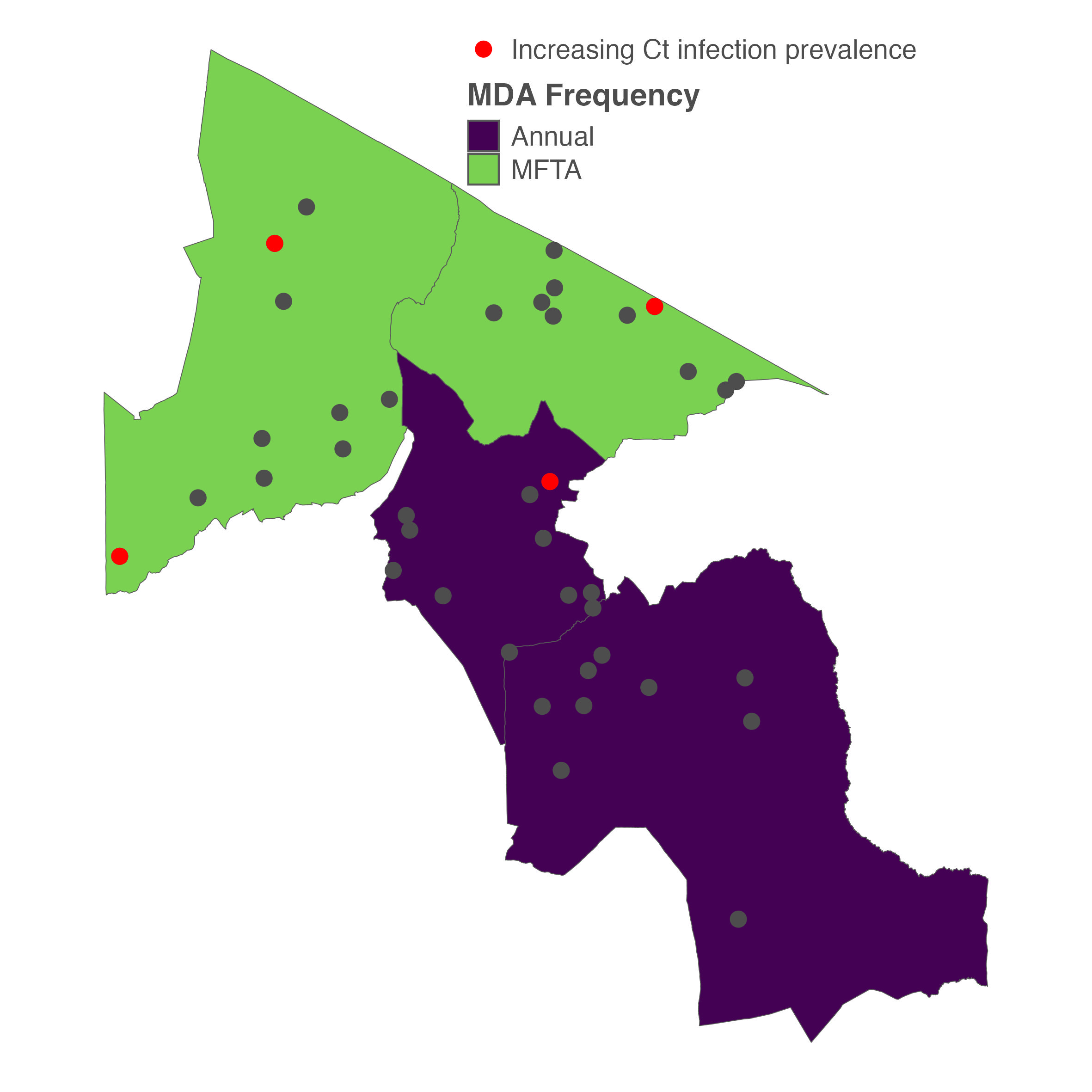

**Figure S3. Map of sentinel sites with increasing ocular Chlamydia trachomatis infection prevalence.**
